# Multimodal Activity-Affinity Assay of ADAM-10 Extracellular Vesicles in Untreated Plasma Reveals Metastatic Stage of Colorectal Cancer

**DOI:** 10.1101/2025.10.17.25338249

**Authors:** Tiger Shi, J. Alex Sinclair, Youwen Zhang, Xuemin Lu, Sonu Kumar, Xin Lu, Satyajyoti Senapati, Hsueh-Chia Chang

## Abstract

Metalloproteinases (MPs) such as a-disintegrin and metalloproteinase-10 (ADAM-10) are key drivers of extracellular matrix remodeling during tumor progression, yet MP-based liquid biopsy tests have not reached clinical utility. Here, we show that active ADAM-10 is selectively enriched on the surface of circulating extracellular vesicles (EVs) in the plasma of colorectal cancer patients. Our findings further suggest ADAM-10+ EVs are locally enriched in dense pre-metastatic tumor extracellular matrix and subsequently accumulate in blood post-metastasis. To capture these unique signatures of disease progression, a novel ADAM-10 activity assay is integrated with a size-selective Immuno-Janus Particle (IJP) affinity assay for characterizing ADAM-10+ EVs in untreated plasma. In a 43-patient colorectal cancer cohort, this multimodal platform distinguished healthy, pre-metastatic, and metastatic states with 95% overall accuracy. When combined with lipidomics as a third modality, the platform correctly determined 97.4% cancer stage accuracy, with only one misclassification. This study establishes a multimodal EV-based activity/affinity assay as a robust framework for liquid biopsy, providing accurate cancer staging, improved prognostics, and offering a potential platform for pan-disease diagnostics.

## 1. Introduction

Colorectal cancer (CRC) remains a leading cause of cancer mortality, with current diagnostics favoring solid tumor detection at advanced stages. Prognostic outlooks decline precipitously following metastasis, with 5-year survival rates falling to 10-20% (Crosby et al., 2022; Guan, 2015; “Survival Rates for Colorectal Cancer,” 2025; “U.S. Cancer Statistics Colorectal Cancer Stat Bite.,” 2025). This presents an unmet need for minimally invasive, repeatable screening techniques that are ideally capable of identifying less entrenched tumors. Liquid biopsy techniques targeting circulating biomolecules and bionanoparticles offer a promising, non-invasive, high-throughput alternative to current screening techniques (Cavallaro et al., 2019; Chen et al., 2020; Kalluri & LeBleu, 2020; Kottorou et al., 2023; Sabatke et al., 2023). However, a liquid biopsy test that can assess the metastatic state has not yet been reported.

Extracellular vesicles (EVs) are a class of lipid-bilayer enclosed nanovesicles secreted by nearly all cell types. They transport proteins, nucleic acids, and metabolites that reflect the molecular state of their parent cell, including cancer-associated changes (Cavallaro et al., 2019; Kalluri & LeBleu, 2020; Kottorou et al., 2023; Li et al., 2023; Sabatke et al., 2023). Tumor-derived EVs circulate abundantly in blood plasma, serving as mediators in tumor progression, metastasis, and microenvironment remodeling (Gao & Li, 2023; Xiong et al., 2020; Zheng et al., 2019). The shift of EV cargo towards pro-tumorigenic inflammatory signaling molecules (Abusamra et al., 2005; Costa-Silva et al., 2015; Fabbri et al., 2012; Shao et al., 2018; Wu et al., 2016) and extracellular matrix (ECM)-modifying enzymes (Moshrefiravasjani et al., 2024; Spugnini et al., 2018; Zhao et al., 2016) makes EVs attractive carriers for established cancer biomarkers. We previously demonstrated that EV-bound proteins such as aEGFR, GPC1, and CEA allow for accurate cancer site identification with AUC values of 0.92-0.99 (Kumar et al., 2025).

One approach for assessing metastatic state relies on preferential release of large EV (*lEV*) over small EV (*sEV*). *sEVs* exhibit CD9, CD63, and CD81 tetraspanin markers. Large extracellular vesicles (*lEVs*) are usually defined as EVs larger than 150 nm, although size overlap with *sEV* is expected. They carry a distinct set of disease markers different from the *sEV*s, due to their direct budding biogenesis instead of the endosomal pathway of *sEV*s biogenesis. Two distinct classes of *lEV*s have recently been identified: microvesicles (MV) and midbody remnants (MBR) (Suwakulsiri et al., 2024). They share some markers with *sEV*s but also carry different markers. The *lEV* to *sEV* ratio has been found to increase in cancer, particularly in metastatic cancer (Park et al., 2023), in senescent cells, and in the plasma of elderly subjects (Alique et al., 2017). However, unlike *sEV*, MV, and MBR-specific *lEV* markers are poorly established, and independent *lEV* immunoassays are not available. Consequently, a rapid diagnostic test that can detect the upregulation of *lEVs* has the potential for detecting cancer metastasis.

Another EV-associated class of biomarkers, metalloproteinases (MPs) are of particular interest. These redox-reactive enzymes regulate ECM remodeling and are linked to tumor growth and inflammation (Cai et al., 2022; Clark et al., 2008; Cox, 2021). A-disintegrin and metalloproteinase-10 (ADAM-10) is a membrane-affixed MP, which is transferred to EVs where they promote vesicle biogenesis, ECM degradation, and migratory signaling, facilitating cancer progression (Aljohmani et al., 2022; Hakulinen et al., 2008; Keller et al., 2020). While soluble forms of ADAM-10 are abundant in plasma, they are less active and not relevant to metastasis as EV-bound MPs (Pelegrini et al., 2025). Inactive ADAM-10 is depleted from the cell surface (Seifert et al., 2021), while EV-bound ADAM-10 remains largely active, situating it as a functional biomarker for mapping cancer evolution. However, existing assays measure either total activity or total protein content. Enzymatic reactions relay key physiological behavior, but are sensitive to sample conditions, and require extensive isolation in addition to a well-designed reporter substrate (Fang et al., 2021; Zhang et al., 2020). Conversely, affinity assays report total ADAM-10, but are blind to enzymatic state and EV association, often lacking the sensitivity to detect EV subsets (Maniya et al., 2024). Thus, no current approach provides EV-specific activity readout of ADAM-10 in plasma. ADAM-10 becomes dysregulated in numerous afflictions, wound healing, and even neurodegenerative diseases (Khezri et al., 2023), so ADAM-10 assays alone cannot screen cancer. They can, however, complement other EV biomarker assays, such as ours (Cheng et al., 2025; Kumar et al., 2024; Kumar et al., 2025; Maniya et al., 2024; Safavi-Sohi et al., 2025; Sharma et al., 2025; Sharma et al., 2023), or serve as a prognosis or therapeutic efficacy assay for cancer patients with known cancer.

In this study, we present an EV-normalized activity-affinity assay of ADAM-10 with a novel Immuno-Janus Particle (IJP) platform (Kumar et al., 2025), which is capable of quantifying relative ADAM-10+ EV abundance and its associated activity directly in plasma. Using progressive stages of CRC patient plasma cohort and four prostate cancer cell lines, we show that ADAM-10 activity originates predominantly from EVs and that the abundance, size, and activity of ADAM-10 EVs differ considerably between pre-metastatic and metastatic patients. To enhance diagnostic performance, we implemented lipidomic analysis using Nile Red lipid dye. We find that the bimodal approach achieved 95% overall metastatic potential accuracy and 89.4% staging accuracy, while the trimodal approach achieved 100% overall metastatic potential accuracy and 97.4% staging accuracy.

These findings establish EV-bound ADAM-10 activity as a biomarker for disease progression and demonstrate that multimodal activity-affinity EV assays can achieve clinically applicable staging accuracy for advancing cancer prognosis and therapy management. Integrating ADAM-10 with a large EV biomarker library could form the groundwork for a massively multi-modal assay and comprehensive pan-cancer liquid biopsy platform.

## 2. Materials and Methods

### 2.1. Clinical Patient Plasma Pre-processing

Operations utilizing patient plasma samples were performed in a BSL-2 biohazard hood. Raw plasma samples were diluted by 10x in 1X DPBS (Gibco, Cat. 14200-075) upon receipt before filtering through MiniSart High Flow 0.22-um (Sartorius, Cat. 16541-K) syringe filters. Samples were aliquoted and stored in-80°C until needed.

### 2.2. Immuno-Janus Particle Fabrication

Immuno-Janus Particles were synthesized in-house. 200 𝜇L of 1.0 𝜇m FluoSpheres™ Polystyrene Microspheres (Thermo Fisher, Cat. F13081/F13083) are added to 10 mL of 70% v/v isopropyl alcohol (VWR, Cat. BDH7999-4). An Electro-Technic Products Inc Model BD-20 High Frequency Generator plasma-treated plain microscope slide (VWR, Cat. 48300-026) for 15 seconds before adding 1 mL of dilute FluoSpheres™ solution on top. The organic solution was allowed to evaporate overnight, leaving a monolayer of dry plain beads. Eight slides were inserted into an AIRCO Temescal FC 1800 electron beam vacuum deposition/thin-film coater system, and coated with 2 nm titanium, followed by 28 nm gold at a deposition rate between 0.5 and 1.0 Å/s. Slides were unaffixed from the PVD machine, and snapped in half longways before being placed in a 1% v/v Tween-20 (Sigma-Aldrich, Cat. P9416-50ML) in DI H_2_O solution. This solution was bath sonicated for 10 minutes in an ultrasonic cleaner (Bransonic^®^, Model 5510R-DTH) to dislodge particles from the slide. The slides were removed from the solution, and the remaining solution was filtered twice with a 5 μm disk filter, Cytiva Whataman™ Puradisc™, Cat. 10463533) to remove aggregates, glass shards, and impurities. The permeate was centrifuged on an AccuSpin Micro 17 (Fisher, Cat. 13100675) at 6000g for 4 minutes before removing the supernatant and concentrating the beads into a volume of about 1 mL, resulting in a final IJP concentration of 1× 10^7^particles/mL. The solution was stored in a 4°C fridge until antibody functionalization for experiments.

### 2.3. Immuno-Janus Particle Antibody Functionalization onto Gold Hemisphere

Gold Conjugation Kit (Abcam, Cat. ab154873) was used to functionalize anti-ADAM-10 monoclonal antibody (Proteintech, Cat. 66620-1-Ig, Lot. 10007605) onto the IJPs’ golden hemisphere. 20 μL of the 1500 𝜇𝑔/mL antibody was combined with 84 𝜇L of Abcam Gold conjugation buffer. 90 𝜇L of this solution was added to 200 𝜇L prepared IJP solution and allowed to incubate for 15 minutes on a shaker (Scilogex, Model MX-M) at 1000 rpm. Abcam Gold conjugation quencher (10 𝜇L) was added to this solution and continued to shake on the shaker for another 15 minutes. The solution was centrifuged in a Fisher AccuSpin Micro 17 at 6000g for 4 minutes and washed with 1:400 Tween-20 (Sigma-Aldrich P9416-50ML): DI H_2_0 once, followed by two washes with 10 times diluted Dulbecco’s Phosphate Buffered Saline (VWR, Catalog no. 02-0119-1000) in DI H_2_O to remove any non-bound antibodies. Functionalized IJPs were resuspended in 50 μL of 10x diluted PBS.

### 2.4. Imaging the Immuno-Janus Particles

The prepared IJP solution was incubated in a 2:1 ratio with each plasma sample in a 0.2 mL PCR tube (Axygen, Ref PCR-02-C). These PCR tubes were placed on a shaker (Scilogex MX-M) at 750 rpm for one hour before imaging. A custom-made apparatus was constructed to image samples effectively. Three cover micro covers (VWR, Catalog no. 48366-089) are stacked and affixed on each side of a microscope slide (VWR, Catalog no. 48300-026). A 2 𝜇L droplet of IJP-sample solution is placed in the middle of the slide, where another microscope cover is placed on top, creating a capillary bridge that suspends the droplet in place. This custom apparatus is placed on an Olympus IX-71 inverted microscope with an Olympus Optical Co. Ltd. 100 W High Pressure Mercury Burner (Model no. BH2-RFL-T3, no. 2308002) supplying the fluorescent signal. The 10x objective was focused 200 𝜇m above the glass slide for recording. Three technical replicates were recorded using a Basler ace 2 R (Basler, Catalog no. a2A1920-160ucPRO) camera at a frame rate of 10 Hz at a duration of 120 seconds per replicate.

### 2.5. Nanoparticle Tracking Analysis Procedure

NTA measurements were performed using NanoSight NS300 (Malvern, UK) (Harper Cancer Research Institute, University of Notre Dame, Notre Dame, IN) per manufacturer protocol (NanoSight NS300 UserManual, MAN0541-01-EN-00, 2017). Each sample was measured via a three-point serial dilution in 1X DPBS to ensure reliability. Camera settings are Capture Gain = 8, Camera Level = 10, Focus = 450, Process Gain = 10, and Detection Threshold = 3. Reported concentrations are scaled to original sample volumes.

### 2.6. ADAM10 Bulk Activity Assay Procedure

ADAM10-specific activity substrate sequence (N to C) consists of a DABCYL quencher, a specific peptide sequence (PRAEALKGGK)(Chen et al., 2021), and a 5-FAM fluorophore. Custom peptide sequence produced via Biomatik Corporation Custom Peptide Synthesis at 90.91% purity and MW 1935.79 Da (Biomatik, Lots. GT91736-SP230343, P240218-LL1143224).

Lyophilized peptide substrate is resuspended in 1X DPBS and solubilized via bath sonication for 1 minute (QSonica, Model Q125 & CL-18). Excitation and emission frequencies for the peptide fluorophore were optimized at 485 nm and 525 nm, respectively (Supporting Fig. 2c). Fluorescent intensities were measured using Tecan Infinite® 200 PRO (Tecan Trading AG, Mannedorf, Switzerland) plate reader. Fluorescent growth rate at limit of detection (LoD) was determined through calibration performed using recombinant human ADAM10 (rhADAM10) (R&D Systems, Cat. 936-AD-020). The calibration curve is shown in Supporting Fig. 2b.

For Michaelis-Menton normalization testing, eight dilutions of ADAM10 substrate (5 - 1000 μM) were formulated and tested against three dilutions (1X, 2D, 4D) of filtered healthy human plasma and filtered healthy human dermal cell cultured media. For cell culture & clinical plasma samples, ADAM10 substrate concentration is 200-μM and samples are tested at 1X concentration and 2D dilution. Each reaction consists of 50-μL substrate and 50-μL sample. Samples were kinetically measured for 120 minutes at 10-minute intervals. Fluorescent growth rates were calculated by calculating slopes for fluorescent intensities measured from 20 to 60 minutes after reaction initiation.

### 2.7. Healthy Human Plasma and Dermal Cell Cultured Media Fractioning and EV Lysis

Diluted and 0.22-μm syringe filtered plasma and cell culture samples were spin filtered using NanoSep 300K Omega (Pall, Cat. OD300C34) spin filters at 2.5kxg for 20 minutes. Flowthrough was reserved, and the retentates were further washed with 1X DPBS. Fractions of the retentate are incubated in a 0.1% final concentration of Triton X-100 (Thermo Fisher, Cat. J62289.AP, Lot. M24J506) for 5 minutes on an orbital shaker at room temperature. Control fractions were taken and incubated simultaneously in equivalent volumes of 1X DPBS. Incubated samples were filtered using SpinX UF500 10k MWCO PES Filter (Corning, Cat. 431478, Lot. 16323005) at 12kxg for 10 minutes and washed using 1X DPBS. All reserved fractions were reconstituted to the original sample volumes.

### 2.8. Total CD63 & ADAM10 ELISA for Healthy Human Plasma Fractions

All samples were evaluated using Human CD63 ELISA Kit (AbCam, Cat. ab275099, Lot. 2101053719) and Human ADAM10 ELISA Kit (AbCam, Cat. ab309315, Lot. 2101052036) per manufacturer protocols. Endpoint fluorescent intensities were measured using Tecan Infinite® 200 PRO (Tecan Trading AG, Mannedorf, Switzerland) plate reader. Calibration curves for CD63 and ADAM10 assays are shown in Supporting Figs. 1c and 1b, respectively.

**Fig. 1.**
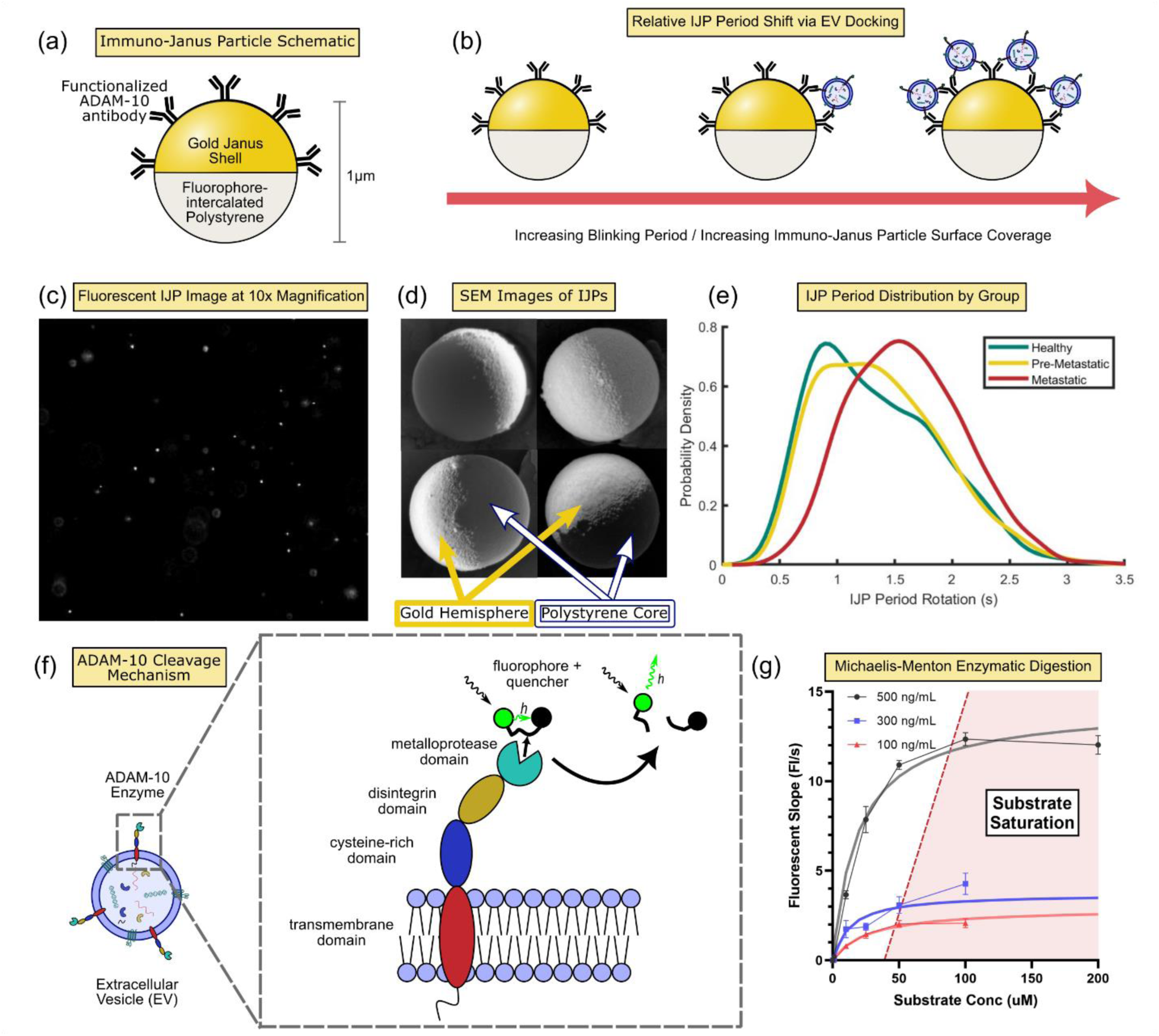
Overview of Prominent Assays for ADAM-10+ EVs (a) Schematic of the Immuno-Janus Particle (IJP) with a fluorescent polystyrene core and gold-coated hemisphere, enabling anisotropic emission. (b) Representation illustrating increased EV docking resulting in longer IJP periods. (c) Video snapshot of IJPs in solution, showing brightness discrepancies from variable particle orientations. (d) Scanning Electron Microscopy images showing IJPs with backscatter electrons; the brighter region is attributable to gold’s higher atomic number, and the uncoated polystyrene is darker. (e) Probability distribution function of anti-ADAM-10 IJP rotational periods following incubation with plasma from healthy, pre-metastatic, and metastatic patients. A rightward shift in period correlates to disease stage. (f) Schematic of membrane-bound, enzymatically active ADAM-10 on an EV surface digesting a synthetic peptide-based molecular beacon. (g) Michaelis-Menten plot of beacon digestion using variable concentrations of recombinant human ADAM-10 (rhADAM-10), yielding fluorescence over time.

**Fig. 2.**
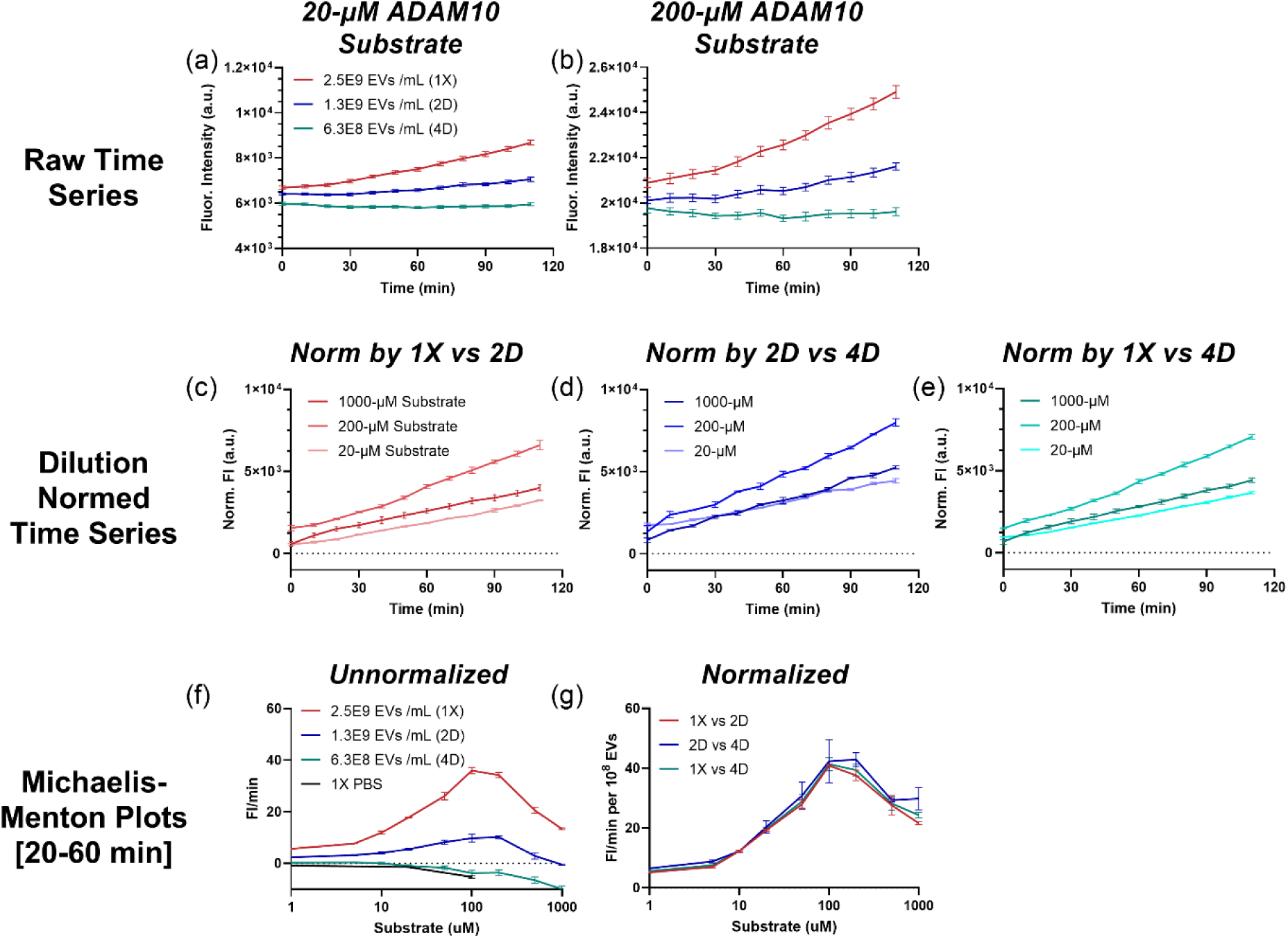
Dilution normalization of bulk activity assay fluorescent evolution time series of EV-associated ADAM-10. (a-b) Fluorescent signal evolution curves for activity-based assays at varying concentrations of ADAM-10+ EVs derived from healthy human plasma samples for (a) 20-μM and (b) 200-μM of ADAM-10 specific activity substrate. (c-d) Internally normalized fluorescent evolution curves using serial dilutions of samples via Equation (1) for various applied concentrations of ADAM-10 specific activity substrates, where the samples are normalized (c) 1x to 2D dilution, (d) 2D to 4D dilution, and (e) 1x to 4D dilution. (f-g) Michaelis-Menten plots for fitted slopes of time series data of (f) unnormalized and (g) normalized curves. Slopes are fitted for time series data at 20-60 minutes after the start of the experiment at 10-minute time steps. All error bars represent measured or propagated standard errors of means.

### 2.9. Prostate Cancer Cell Cultured Media and EV Collection

PC3, DU145, 22Rv1, and LNCaP prostate cancer cell lines were maintained in 10 cm culture dishes in RPMI-1640 medium (Gibco) supplemented with 10% fetal bovine serum (FBS; Gibco). Upon reaching confluence, cells were washed twice with phosphate-buffered saline (PBS; 1×) to remove residual serum components. The culture medium was then replaced with 10 mL of RPMI-1640 containing 10% exosome-depleted FBS (Gibco), and cells were incubated for 48 h. Conditioned media were subsequently collected for exosome isolation.

### 2.10. Bradford Total Protein Assay Procedure

Total protein assay was designed using 150-μL Bradford Reagent (Sigma Millipore, Cat. B6916-500ML; Lot. SLCN3585) applied to 5-μL of each target sample. Samples are incubated for 30 minutes on an orbital shaker at room temperature. Absorbance intensities at 595 nm were measured using Tecan Infinite® 200 PRO (Tecan Trading AG, Mannedorf, Switzerland) plate reader. Protein standards (1 – 2250 μg/mL) were formulated from step dilutions of Bovine Serum Albumin 30% in DPBS (Sigma Millipore, Cat. A9576-50mL, Lot. SLCL8813). The calibration curve for the Bradford total protein assay is shown in Supporting Fig. 3c.

**Fig. 3.**
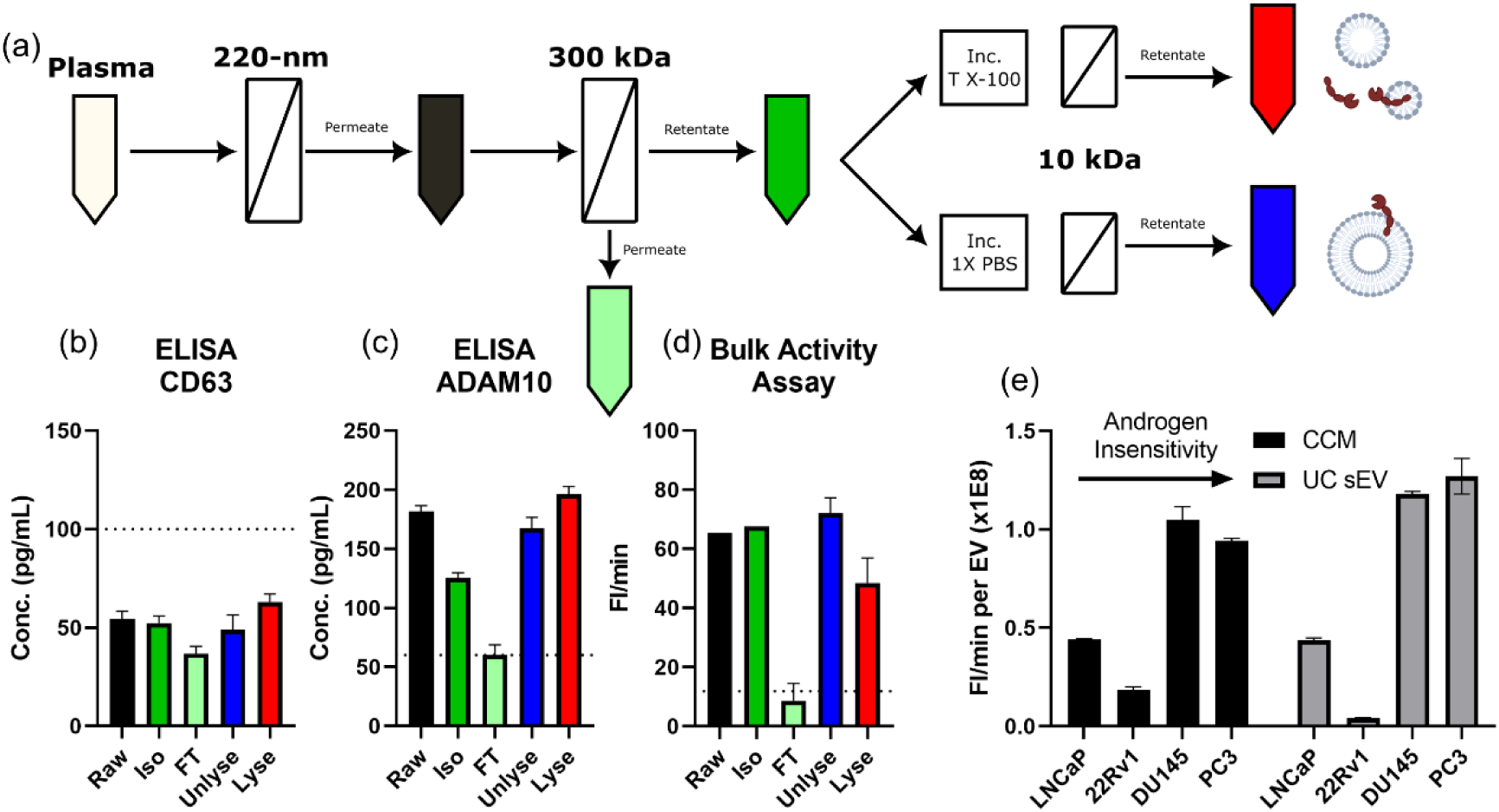
ELISA and bulk activity assay of healthy human plasma fractions and PCa-derived cell lines. (a) Sequential filtration workflow of healthy human plasma. Samples filtered through a 220 nm syringe filter and a 300 kDa filter. The 300 kDa retentate (vesicle-rich fraction) was either lysed with 1% Triton X-100 or washed with PBS before removing small components with a 10 kDA filter. Colors shown here are maintained for subsequent panels. (b) CD63 ELISA quantification shows low signal across all five plasma fractions. (c) ADAM-10 ELISA quantification shows detectable signals for all plasma fractions except the 300 kDa permeate (vesicle-free fraction), suggesting vesicle-associated quantification. (d) Bulk ADAM-10 activity assay mirrors trends found in ADAM-10 ELISA for the five plasma fractions, with low activity for the 300 kDa filter permeate, and high signal in vesicle-containing fractions. (e) Normalized ADAM-10 activity per 10^8^ EVs for four prostate cancer cell lines of increasing insensitivity to androgen: LNCaP, 22Rv1, DU145, and PC3. Higher androgen insensitivity in both cell culture media and isolated EVs suggests higher enzymatic activity correlates to disease progression.

### 2.11. Nile Red Total Lipid Assay Procedure

Lyophilized Nile Red (Thermo Fisher, Cat. N1142, Lot. 2916406) is reconstituted to 1 mg/mL in 1X DPBS. Samples are incubated at a 20-μg/mL final concentration of Nile Red for 30 minutes before imaging. Excitation and emission frequencies for the Nile Red assay were optimized at 555 nm and 635 nm, respectively (Supporting Fig. 3b). Fluorescent intensities were measured using Tecan Infinite® 200 PRO (Tecan Trading AG, Mannedorf, Switzerland) plate reader. The calibration curve for the Nile Red total lipid assay is shown in Supporting Fig. 3a.

### 2.12. Unsupervised Learning Procedure for Principal Component Analysis

Group expressions were compiled for all 43 patients whose samples were analyzed in this study. An average and standard deviation were calculated for the healthy group for each of the six studied parameters. All samples were subtracted from the average healthy expression and were then divided by the healthy standard deviation to determine a z-score. MATLAB’s pca function was used to calculate and store the principal component coefficient matrix, variable weights, and loadings, as well as the variance explained by said parameter. These values were used to determine parameters contributing most significantly towards the system variance and the degree attributed.

### 2.13. Statistical Analysis

The data presented here is expressed as the mean and standard deviation or standard error of the mean as calculated using Excel, Python, or GraphPad Prism 8 software functions. Errors are propagated through formulas using standard propagation of uncertainties techniques.

## 3. Results and discussion

### 3.1. Immuno-Janus Particles Capture and Report ADAM-10+ Vesicles

To quantify ADAM-10+ EVs, we implement our novel Immuno-Janus Particle (IJP)(Kumar et al., 2025) sensor. The IJPs are 1000-nm fluorescent polystyrene beads coated with a 30 nm golden hemisphere to quench the fluorescent signal when viewed orthogonally (Fig. 1a,d). The gold surface can facilely conjugate a variety of antibodies, including 𝛼-ADAM-10, as represented in Fig. 1a,b. When these particles undergo chaotic thermal fluctuations in solution, they exhibit an observable blinking phenomenon (Fig. 1c). The blinking frequency is inversely related to particle size, represented by the rotational diffusivity 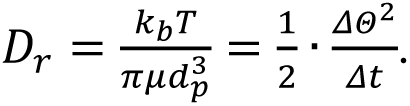 The-3 scaling of the rotational diffusivity, in contrast to the-1 scaling for translational diffusivity, renders the blinking frequency very sensitive to the particle size. This size sensitivity not only provides precise quantification of the effective size increase due to EV binding but also reduces noise from dispersed ADAM-10 with a much smaller size than EVs. By observing the relative blink rates, we can approximate the effective size of each tracked particle. Irreversible EV-IJP binding expands the hydrodynamic particle diameter with the number of EVs and the size of the bound EVs. The blinking frequency of IJP is hence a sensitive and selective metric for ADAM-10+ EV number and size (Fig. 1b,e). Since *lEV* is only 10% larger than *sEV*, our detected 50% difference in IJP blinking period is most likely due to a change in the number of ADAM-10+ *lEVs*.

A custom single-particle tracking software captures the blinking dynamics of each detected IJP over the course of a 60 second recording (Fig. 1c). Ensemble average particle time periods are extracted using individual continuous wavelet transformations. Fig. 1d shows the SEM images of the hemisphere gold-coated Janus particles. Fig. 1e shows probability distribution functions of the three metastatic stages. Each curve represents the cumulation of every IJP period measurement for all individual patients within their respective group across the entire study. This constitutes thousands of IJPs demonstrating the increasing upregulation of ADAM-10+ EVs throughout CRC progression, with metastatic cancers having a noticeable increase in IJP blinking period.

### 3.2. Off-target Interference of ADAM-10 Activity Can Be Normalized Through Serial Dilution

To assess total enzymatic activity, we adapted a digestible fluorophore peptide beacon based on the (PRAEALKGGK) sequence from prior work (Chen et al., 2021). The beacon consists of a 5-FAM fluorophore and a peptide-adjacent DABCYL quencher, so that the proteolytic cleavage emits a measurable fluorescent signal, as shown in Fig. 1f. Soluble and membrane-bound ADAM-10 are both capable of cleaving this substrate. This fluorescent growth over time is determined by classical Michaelis-Menten kinetics, defined by the rate law 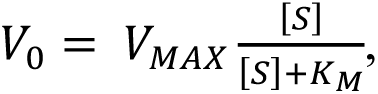 where 𝑉0 is the initial reaction rate, 𝑉_𝑀𝐴𝑋_ = 𝑘_𝑐𝑎𝑡_ · ([𝐸] + [𝐸𝑆]) = 𝑘_𝑐𝑎𝑡_ · [𝐸]_0_ is the maximum reaction rate, and [𝑆] is the peptide substrate concentration. 𝐾_𝑀_ is the Michaelis constant, defined by 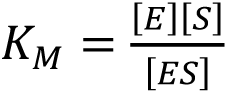 [𝐸] and [𝐸𝑆] being enzyme and enzyme-substrate complex concentrations, respectively (Van Daela, 2015). At a high applied concentration of substrate [S], the rate equation simplifies to 𝑉_0_ = 𝑉_𝑀𝑎𝑥_ = 𝑘_𝑐𝑎𝑡_ · [𝐸]_0_ and is linear with respect to the total concentration of enzymes in the system. Using three concentrations of recombinant human ADAM-10 (rhADAM-10) in Fig. 1g, a substrate concentration of 100 μM was selected as optimal since it lies wholly within this saturation region, allowing us to attribute any changes in reaction rate solely to differences in enzyme concentration.

When subjected to human plasma, the fluorescent time series (Fig. 2a,b) appear non-linear until after an hour of reaction initiation. With this skewed data, the resultant Michaelis-Menton plots (Fig. 2e) appear non-standard. This is due to interfering agents within each sample. However, by running each sample in replicates of serial dilution, the underlying linear enzymatic activity rates can be uncovered by applying the normalization function 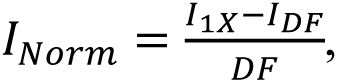 where *DF* is the dilution factor, yielding linearity within the first hour of reaction (Fig. 2c,d). The Michaelis-Menton plot featuring normalized reaction rate for a variety of different levels of dilution (Fig. 2f) demonstrates the convergence of each normalization to the underlying reaction rate while removing interfering noise from the sample. Using this methodology, subsequent activity assays can be internally normalized without extensive sample processing and cleaning.

### 3.3. ADAM-10+ Vesicles Enhance Enzymatic Activity

To test the dependence of ADAM-10 activity on its location on the EV membrane, the workflow illustrated in Fig. 3a was implemented. Healthy human plasma was filtered through a 220 nm filter to remove cell debris and large vesicles. The permeate (Raw) is passed through a 300 kDa centrifugal filter, where the twice-filtered permeate (FT) contains the free-floating protein fraction, while the retentate (Iso) retains the plasma vesicles (<220 nm) such as EVs. The EV isolate (Iso) was reconstituted to the original sample volume using 1X PBS and split into two equal 400-μL fractions: (1) one sample was given 20-µL of 2% Triton X-100 surfactant (Lyse), and (2) the other control sample received 20-µL of 1X PBS (Unlyse). Both samples were incubated for 30 minutes at room temperature on an orbital shaker and subsequently washed in a 10-kDa centrifugal spin filter with three resuspensions of 400-µL 1X PBS to remove remaining surfactant. The samples were each resuspended in 400-µL of 1X PBS. All samples (Raw, Iso, FT, Unlyse, & Lyse) were assayed using total CD63 ELISA, total ADAM-10 ELISA, and bulk ADAM-10 activity assay.

Looking at the ELISA results (Fig. 3b), we find that CD63 was too low in concentration in all samples to be quantified. At the Raw and Unlyse samples, the ADAM-10 concentration and activity were retained in the EV through the 300 kDa and 10 kDa spin filtering steps (Fig. 3c,d). Along with this, despite the apparent drop in ADAM-10 concentration for the Iso sample, the activity rate is retained. This behavior suggests that the drop is likely due to aggregation of nano-carriers in the spin-filtering rather than a true loss in enzyme concentration. Additionally, we notice that for both ADAM-10 concentration and activity rate, the FT fraction containing the free-floating proteins holds no detectable level of ADAM-10 presence or activity (Fig. 3c,d). Ultimately, upon lysis of the nano-carriers in the sample, we notice a 10% increase in ADAM-10 concentration and a simultaneous 35% reduction in ADAM-10 activity rate. The above results suggest that ADAM-10 in healthy plasma is primarily located on the surface of nano-carriers such as EVs. Furthermore, delipidation of ADAM-10+ EVs releases a small additional amount of ADAM-10 into solution (Fig. 3c), yet this causes a significant reduction in enzymatic activity (Fig. 3d). These findings suggest that ADAM-10 activity is enhanced when embedded in a lipid membrane, likely due to the anchored domain securing the enzyme’s structure (Pelegrini et al., 2025). Disrupting this membrane system increases the amount of solubilized protein, as the internal cargo is jettisoned into the bulk. However, this disruption destabilizes the membrane proteins, leading to reduced activity. As such, the ADAM-10 species of highest enzymatic function, and the ideal species to analyze from hereon, are intact EVs. These trends were verified for 220 nm filtered cell-cultured media (CCM) produced by patient-derived healthy human dermal cells as well (Supporting Fig. 1). These cells were shown to inherently produce high ADAM-10 activity on their EVs.

### 3.4. ADAM-10 Signal is Expressed Primarily on EVs and Correlates with Metastatic State in Prostate Cancer Cell Cultures

ADAM-10 *in vitro* studies link its overexpression to increased metastasis in colon, gastric, prostate, ovarian, uterine, and leukemia cancer cell cultures (Saha et al., 2023), underscoring its entanglement with cancer progression. Subsequently, we applied this enzymatic activity assay to prostate cancer (PCa) cell lines of varying metastatic potential, including LNCaP, 22Rv1, DU145, and PC3, evaluating both cell culture media and ultracentrifugation (UC) isolated EVs (Fig. 3e). Here, we used androgen sensitivity as a proxy metric for disease progression, where prostate cancer cells become more insensitive to androgen deprivation therapy as the disease progresses towards later stages. A general trend emerged where androgen-insensitive cell lines exhibit a higher EV-normalized enzymatic activity, with the exception of 22Rv1, showing markedly lower enzymatic digestion. Akin to the plasma results, enzymatic activity rates were consistent across both unprocessed media and isolated EVs. These findings support a model where the enzymatically relevant ADAM-10 is confined to EV membranes. Pairing the IJP platform, which is independently capable of quantifying ADAM-10+ EVs, allows us to gain a dual-modality screening technique. One test quantifies vesicle abundance and the other quantifies enzymatic capacity, offering dynamic insight into cancer-related vesicle heterogeneity through the oncogenic process. As shown in Fig. 3d, the raw plasma filtered through a 220 nm membrane produces comparable enzymatic signals to higher purity vesicle fractions (300 kDa retentate and 10 kDa PBS washed retentate). Combining this fact with the IJP platform’s immunity to soluble protein interference, this motivates our use of minimally processed plasma in our subsequent EV-centric activity/affinity analyses. We implement both enzymatic and IJP assays in a pilot study of 43 patient sample cohorts, alongside additional orthogonal characterization.

### 3.5. Unsupervised Learning to Determine Parameters with Diagnostic Efficacy

The comprehensive plasma analysis consists of six orthogonal measurements: bulk ADAM-10 enzymatic activity, Immuno-Janus Particle rotational period to measure ADAM-10+ extracellular vesicle concentration, NTA-derived particle concentration, NTA-derived mean particle diameter, plasma lipid content quantified by Nile Red staining, and total protein concentration measured by Bradford assay.

Because these metrics span disparate numerical scales (e.g., IJP period: 1.16-1.73 s, and nanovesicle concentration: 3.51 × 10^10^ − 2.44 × 10^12^ particles/mL), raw values were converted to Z-scores relative to the healthy cohort mean values:

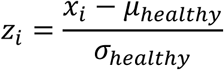

Group-averaged Z-scores are shown in Fig. 4a, and individual patient expression in Fig. 4b. We observe a single metastatic outlier (M7) whose expression levels are inconsistent among group behavior for most analyzed parameters, save the IJP period. The sample’s aberrant viscosity, color, and turbidity suggest likely hemolysis during plasma preparation (Sowemimo-Coker, 2002). Across disease progression from healthy to metastatic states, we observe increased IJP period, particle size, and protein concentration, contrasted against a decreased ADAM-10 activity, particle concentration, and lipid content. These trends are not strictly linear; for example, total protein concentration plateaus in pre-metastatic cancer, before declining in metastatic patients. Non-linearity suggests attributes are highly specific to their respective stage.

**Fig. 4.**
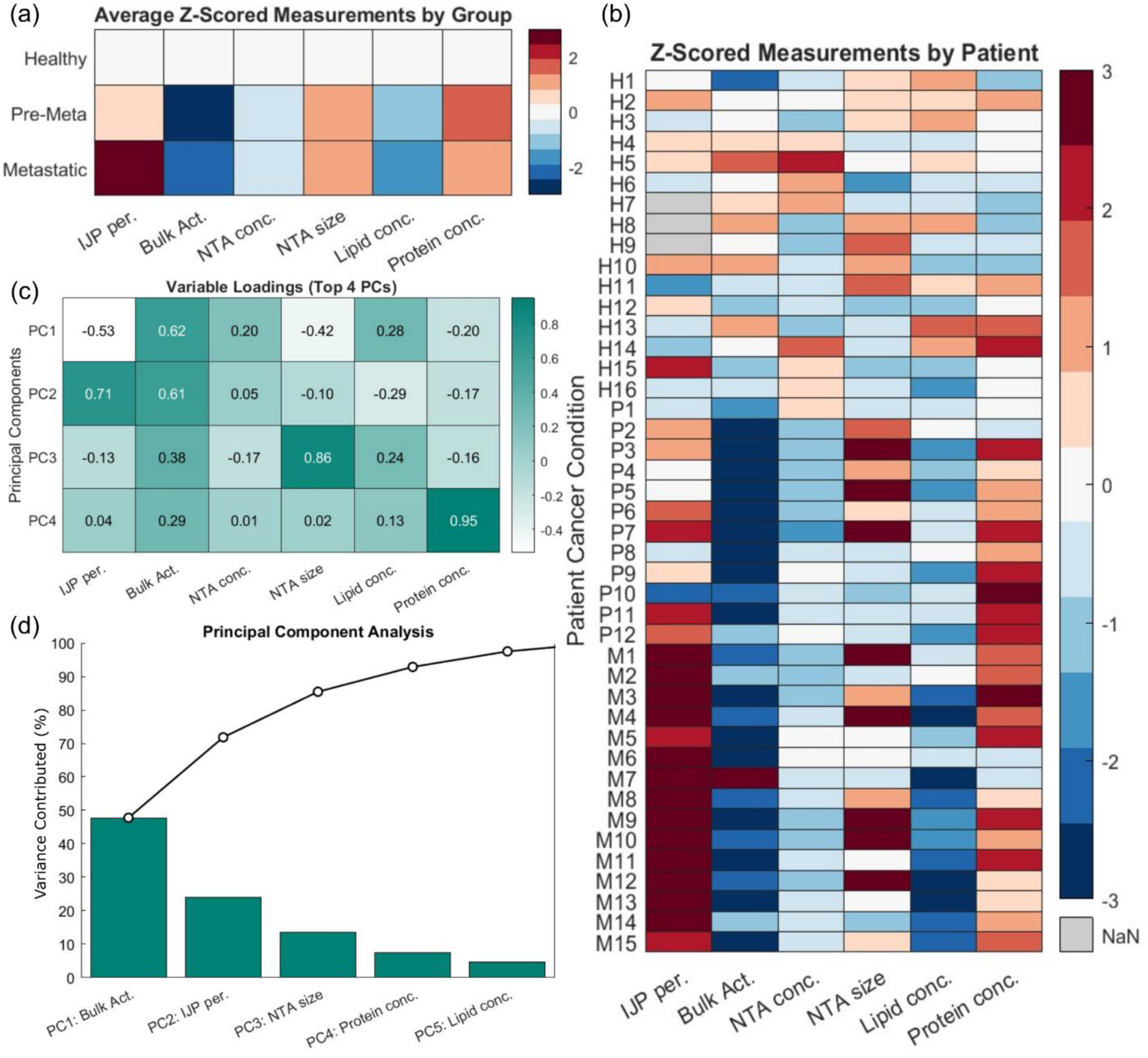
Multimodal plasma profiling and diagnostic classification using Z-score normalizations and principal component analysis. (a) Heatmap of group-averaged Z-scores for six parameters: Immuno-Janus Particle (IJP) rotational period, bulk ADAM-10 activity, nanoparticle tracking analysis (NTA) particle concentration, NTA mean particle diameter, plasma lipid content (Nile Red fluorescence), and total protein content. Values normalized to the healthy cohort mean. (b) Heatmap of individual patient Z-scores; gray cells indicate unmeasured parameters. (c) Variable loading heatmap showing the contributions of each parameter to the top four principal components (PCs) derived from PCA. Color grading reflects the magnitude and direction of loadings. (d) Pareto plot showing variance explained by each PCs and their respective top-loading variable.

To identify parameters with the greatest diagnostic potential, we performed unsupervised Principal Component Analysis (PCA) (Abdi, 2010). PCA projects interdependent correlated variables onto orthogonal Principal Components (PCs) ranked by degree of variance explained. Variable loadings indicate the relative influence of the six original measurements on an individual PC, with higher absolute values representing a stronger contribution. In Fig. 4c, bulk ADAM-10 enzymatic activity contributes the highest to PC1, with a loading of 0.62, while the IJP period is highest for PC2 (0.71). These two leading components contribute 47.7% and 24.1% variance, respectively, explaining 71.8% of system variance (Fig. 4d), indicating a bimodal ADAM-10 quantification is likely diagnostically sufficient for CRC stage classification. For additional diagnostic potency, PC3 (mean particle diameter, loading = 0.86) and PC4 (protein concentration, loading = 0.95) can be incorporated, contributing an additional 13.6% and 7.4% of system variance, respectively (Fig. 4c-d). Importantly, this small change in EV diameter from an orthogonal NTA characterization confirms that the change in IJP period is due to a change in ADAM-10+ EV concentration. Our findings from principal component analysis suggest that leveraging ADAM-10 activity and the IJP period would provide a minimalistic, diagnostically accurate screening platform for cancer staging. We will explore this relationship and uncover trends in the next section.

### 3.6. Colorectal Cancer Progression is Reflected in ADAM-10+ Vesicles and Bulk Activity

We explored the diagnostic potential of combined orthogonal IJP period and bulk ADAM-10 activity for colorectal cancer progression. In Fig. 5a, healthy patients exhibit elevated bulk activity and quicker IJP rotation, whereas pre-metastatic samples show a marginally increased IJP period but a significantly reduced bulk activity. Metastatic samples are characterized by significantly larger IJP blinking periods and slightly higher bulk activity, except metastatic patient M7, who shows unusually high ADAM-10 activity despite having a characteristic IJP period. The orthogonal separation between activity and affinity for premetastatic and metastatic samples in Fig. 5a is striking and underscores the need for a bimodal assay for accurate metastatic assessment.

**Fig. 5.**
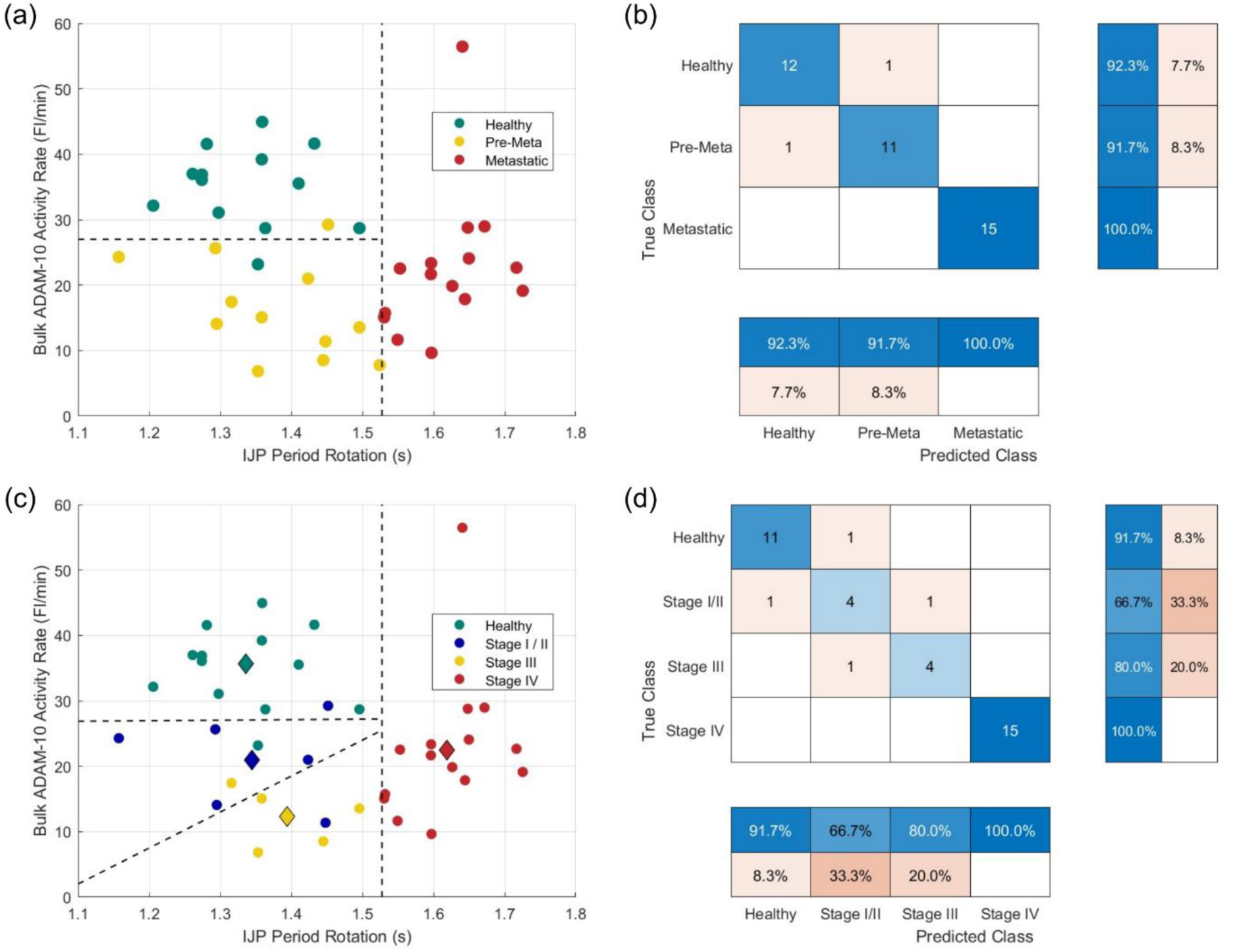
Bimodal analysis of bulk ADAM-10 enzymatic activity rate and Immuno-Janus Particle period enables accurate classification of metastatic potential and colorectal cancer stage. (a) Scatter plot of bulk ADAM-10 activity (y-axis) versus IJP period (x-axis) for individual patients. Dashed lines represent decision boundaries separating healthy, pre-metastatic, and metastatic groups. (b) Confusion matrix for linear discrimination classification showing true labels (rows) versus predicted labels (columns). Group precision and recall are displayed in the right-most and bottom panels, respectively. (c) Scatterplot with refined classification, dividing pre-metastatic samples into Stage I/II and Stage III cancer. Diamonds represent average expressions of their respective group, and circles represent individual patient data. One pre-metastatic sample was excluded as it was not assigned a stage. (d) Confusion matrix showing classification performance for stage-wise analysis using bimodal input variables.

Applying simple thresholding values, the IJP period >1.53s perfectly isolates the metastatic samples. Pairing faster periods with a bulk activity of 27 FI/min defines the pre-metastatic and healthy cohort boundaries, allowing for the implementation of a linear discrimination classifier in Fig. 5b. The precision/recall values achieved are 100% for metastatic, 91.7% for pre-metastatic, and 92.3% for healthy samples across 40 total patients. Misclassifications were limited to one sample in both non-metastatic groups, granting an overall 95% accuracy. As a pilot study, these results justify further investigation into the interdependence of ADAM-10 activity and ADAM-10+ EV content as bimodal disease progression markers. Surprisingly, the fraction of active ADAM-10+ plasma EVs is lower in early premetastatic patients than in healthy samples, while the concentration of both active and inactive ADAM-10+ EVs increases for post-metastatic samples.

To explore the stage resolution of this capture and activity-based study, we partitioned the pre-metastatic cohort into Stage I / II cancer and Stage III cancer at the time of diagnosis (Fig. 5c). Stage I was combined with Stage II due to sample quantity, and one unstaged pre-metastatic sample was omitted. The Diamond symbols in Fig. 5c represent the group stage-wise behavior. It shows that Stage I/II experience an early reduction in enzymatic activity, and a slight increase in ADAM-10+ EV quantity. Stage III continues this same general trend. Stage IV, metastatic samples demonstrate an increase in activity and the largest adjacent inter-group IJP period increase, indicating the most drastic shift in vesicle’s conformational volume upon the initial metastasis of the tumor. Patients of the same stage have similar IJP periods and enzymatic activity, resulting in a tight clustering of the group behavior. Subsequently, a few misclassifications occurred between adjacent stages (i.e., no healthy sample was identified as stage III/IV).

Stage-based identification in Fig. 5d achieved a diagnostic accuracy of 89.7%, with all misclassifications occurring in early-stage samples, adjacent or within Stage I/II. This high staging resolution underscores the merit of incorporating activity-and expression-based diagnostics, accurately pinpointing stage and allowing for earlier, more robust cancer interventions.

### 3.7. ADAM-10 Enzymatic Disparities among Cells, Cell Culture EVs, and Plasma EVs

Healthy patient plasma exhibits the highest ADAM-10 activity despite having the fewest ADAM-10+ EVs as defined by the IJP period (Fig. 5a,c), a trend opposite to that observed in cell-culture assays (Fig. 3e) where metastatic cell lines exhibit elevated activity. This discrepancy likely reflects differences in therapeutic intervention or the culture system tumor physiology. In solid tumor environments, EV transport through the unvascularized extracellular matrix is innately limited, a behavior not captured in monolayer cultures or more advanced organoid models. Moreover, transport resistance is selectively more severe for EVs with active ADAM-10, given their function to bind to cell-surface receptors and initiate cleavage events on the order of hours (Fig. 2). Hence, active ADAM-10 EVs are enriched locally within the tumor microenvironment, while their dissipation into circulation is diminished relative to inactive EVs.

Moreover, regulatory mechanisms not captured in cell culture may further limit plasma ADAM-10 activity in cancer patients. Membrane-bound ADAM-10 enhances cancer proliferation through erbB ligands such as EGF (Moss et al., 2007), but therapeutic inhibition of the EGFR signaling elevates ADAM-10 pro-domain levels and enhances ADAM-10 self-dimerization, collectively suppressing ADAM-10 activity (Deng et al., 2014; Moss et al., 2007; Wetzel et al., 2017). Reduced cholesterol in unvascularized tumors due to low nutrient transport may further impair ADAM-10 function, which mirrors regulatory pathways described for ADAM-17; the metalloproteinase’s proximity to lipid rafts and cholesterol-dependent interactions modulate shedding efficiency (Tellier et al., 2006). We observe a correlated level change in lipoproteins that transport cholesterol (Fig. 6). We suggest the synergistic factors of transport limitation, receptor engagement, metabolic inhibition, and lipid dysregulation, which result in diminished production and circulation of active ADAM-10 EVs in cancer patients, especially the pre-metastatic niche.

**Fig. 6.**
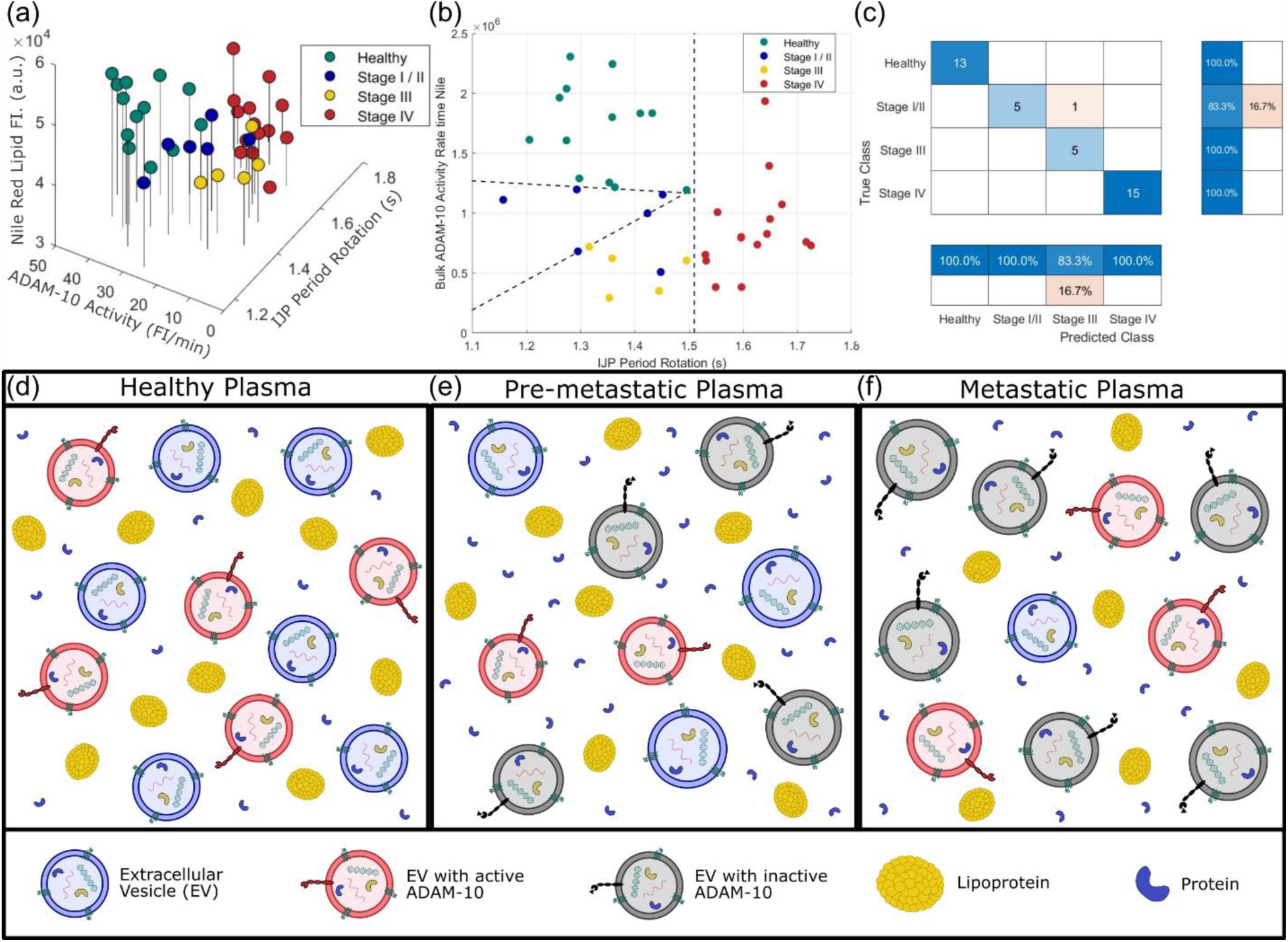
Trimodal analysis of lipid content, enzymatic activity, and IJP period enhances diagnostic staging of colorectal cancer and schematic summary plasma profiles. (a) Ternary plot of IJP period, bulk ADAM-10 activity, and Nile Red lipid fluorescence across healthy, Stage I/II, Stage III, and Stage IV cohorts. (b) Two-dimensional projection showing IJP period versus the multiplication of ADAM-10 activity and lipid content. Dashed lines indicate classification thresholds. (c) Confusion matrix showing 97.4% overall accuracy of stage-wise performance using trimodal analysis. (d-f) Schematics of plasma composition across disease progression, showing an increase in proteins, a decline in protein, and a reconfiguration of ADAM-10 to inactive form, and more abundant forms.

A decrease in plasma EV ADAM-10 activity has been previously noted in metastatic patients (Tugutova et al., 2019), aligning with other reports finding disease-associated dysregulation of metalloproteinase activity (Aljohmani et al., 2022; Ebsen et al., 2013; Moss et al., 2007; Wetzel et al., 2017). These studies, along with our findings, support a spatial model of ADAM-10 regulation. Locally elevated enzymatic activity at the tumor site facilitates ECM remodeling and invasion, whereas distant EV-associated activity is reduced by regulatory constraints, including ECM density, lipid raft disruption, and pro-domain mediated inhibition. This framework reconciles the paradox between elevated ADAM-10 activity in tumors and cell culture, with a reduced EV-normalized activity observed in pre-metastatic plasma. Additionally, in vascularized metastatic tumors, increased EV release, enhanced vascular access, and the more permeable ECM would lead to the observed modest recovery of ADAM-10 activity per EV.

### 3.8. Trends in ADAM-10+ EVs and Composition across Colorectal Cancer Progression

To improve upon the separation of disease cohorts, we posit the merits of a third diagnostic modality, such as NTA size distribution, which contributed to the highest system variance following enzymatic activity and IJP period (Fig. 3c,d). Although metastatic samples display a general positive correlation between particle size and disease state (Fig. 4c), the high intragroup size variability severely constrains diagnostic resolution. Because particle size does not statistically determine cancerous diseases from healthy groups, we sought a modality that consistently downregulates during tumor development. Nile Red lipid signal decreases significantly alongside ADAM-10 activity (Fig. 4b), making it a viable option for analysis.

Fig. 6a presents a ternary analysis merging the IJP period, bulk ADAM-10 activity, and Nile Red lipid fluorescence. The three-dimensional plot of the four groups reveals distinct clustering, offering improved separation compared to IJP rotation and ADAM-10 activity alone. enhanced separation. Due to the inclusion of the Nile Red reported lipid content, a statistically significant decrease in signal, second only to the enzymatic rate, is achieved (Fig. 4b). Notably, both lipid content and vesicle concentration decline from healthy to metastatic stages, suggesting a reduction in lipid-rich lipoproteins(Jiang et al., 2006; Kokoglu et al., 1994; Kritchevsky et al., 1991; van Duijnhoven et al., 2011) or exosomal subtypes of EVs. This association helps reconcile the discrepancy of higher ADAM-10+ EVs concentration but fewer total particles.

A simplified two-dimensional plot (Fig. 6b) demonstrates the power of this three-variable analysis. By multiplying the ADAM-10 activity and the Nile Red lipid content, we achieved a clear separation of the healthy and pre-metastatic samples, enabling an accurate disease staging assessment. The selected delineations achieved 100% accuracy for both cancer presence and metastatic potential, and an impressive 97.4% accuracy for stage determination. Only a single stage I/II patient was misclassified as stage III (Fig. 6c), underscoring the likelihood that large-scale clinical application will inevitably blur the sharp thresholds. Regardless, the strong stage-dependent trends discovered across this 43-patient pilot study hold significant translational potential. Combining the stage-wise diagnosis in this work with the disease type screening from our previous work (Kumar et al., 2025) would allow us to identify cancer location and progression in <60 minutes with a single blood draw.

To visually contextualize the trends found in this multimodal analysis of 43 patients spanning three disease states and six diagnostic parameters, we constructed simplified schematic overviews of plasma composition across metastatic progression (Fig. 6d-f). Relative to healthy expression levels, diseased plasma contains reduced lipoproteins, increased soluble proteins, and a shift towards inactive EV-bound ADAM-10, leading to lengthened IJP periods without a reciprocal increase in bulk enzymatic activity assay. Tumor-associated EV release increases in late-stage disease (Bebelman et al., 2021; Surman et al., 2017; Xu et al., 2018), with higher ADAM-10+ EV concentration coinciding with a substantial reduction in lipid-rich particles. Reduced EV concentration in pre-metastatic samples may result from tumor therapy or tumor-induced metabolic suppression of non-tumor cells, further enhancing the proportional contribution of tumor-derived EVs.

This compositional shift suggests a redistribution of ADAM-10+ EVs to larger vesicles such as *lEVs* or microvesicles. Prior studies report ADAM-10 enrichment in the small EV fractions of (100K ultracentrifugation pellets) (Kowal et al., 2016), yet large EVs (100-1,000 nm) such as microvesicles and ectosomes are increasingly abundant in cancer plasmas (glioblastoma, non-small-cell lung carcinoma, multiple myeloma) and carry pertinent established markers (e.g., MUC1, CEA, CA19-9) (Surman et al., 2017). Our observations of larger particles and slower IJP blinking rates in metastatic plasma support this transition. Expanding multimodal assays to incorporate additional modalities for larger EVs, such as our recent Surface Acoustic Wave sensor platform (Cheng et al., 2025), would discern the various factors regarding ADAM-10 carriers through disease progression. Ultimately, a multimodal affinity-activity assay on colocalized biomarker analysis serves as the most comprehensive, minimally invasive screening assay for cancer detection and staging. To improve clinical applicability, low-dimensional Principal Component Analysis enhanced by AI for large-library activity/affinity assays across a breadth of diseases.

## 4. Conclusion

This work establishes a bimodal activity/affinity assay for ADAM-10+ EVs as a promising platform for assessing the metastatic status in colorectal cancer. We demonstrate that intact plasma (EVs) carrying the enzymatically active forms of ADAM-10, and that these vesicles account for the majority of the signal observed in bulk plasma enzymatic assays. By analyzing progressively metastatic, androgen-insensitive prostate cancer cell lines (LNCaP, 22Rv1, DU145, and PC3), we observe a general increase in ADAM-10 enzymatic rate dependent on the EV concentration.

Multimodal profiling combined with PCA revealed that IJP rotation period (a surrogate for ADAM-10+ EV abundance), combined with bulk ADAM-10 activity, explained a majority of the diagnostic variance. The bimodal screening enabled a 95% accurate metastatic potential identification, while implementing lipidomics as a third modality achieved 100% accuracy for cancer identification and metastatic potential, and 97.4% accuracy for stage classification. These results highlight the importance of integrating multiple affinity/activity-based analytical methods into a unified framework for orthogonal EV liquid biopsies.

Beyond diagnostic performance, our findings ascertained mechanistic insights into ADAM-10 regulation in cancer. Active ADAM-10+ EVs are selectively retained with unvascularized tumors due to transport limitations, protein interactions, and pro-domain mediated regulation. With disease progression, ADAM-10+ redistributes from smaller EVs to larger ectosomes and microvesicles, amplifying the difference in IJP period between premetastatic and metastatic data. Concurrent declines in lipid-rich particles further contribute to the shift in plasma content we observed. These findings underscore ADAM-10-based multimodal EV assays as highly selective tools for prognosis, disease staging, and therapeutic monitoring. Augmentation of established disease markers or MPs, affinity/activity multiplexing, and machine learning analysis of a large library of assays could bolster this platform into a scalable screening platform for a wide breadth of cancers, in addition to other diseases.

## Ethics approval and consent to participate

We received colorectal cancer and healthy group plasma samples from Precision for Medicine, and unlabeled and randomized immediately upon arrival. An approved IRB protocol is already in place at Precision for Medicine for the collection of plasma samples from patients. Precision for Medicine works with regulatory authorities and accrediting organizations around the world to ensure that the sample collection process and protocol follow the latest FDA, EMA, and MHRA guidelines. All procedures performed in studies involving human participants were conducted in accordance with the ethical standards of the University of Notre Dame.

## Availability of data and materials

The datasets used and/or analyzed during the current study are available from the corresponding author on reasonable request.

## Authors’ contributions

H.C.C., S.S., T.S., J.A.S., and S.K. conceptualized the study and designed the experiment. T.S. and Y. Z. designed and conducted the activity assay. T.S. conducted NanoSight, lipidomics, and proteomic experiments. J.A.S. fabricated the IJP and conducted the IJP experiments. Xuemin Lu and Xin Lu provided the metastatic cell culture media and invaluable advice. H.C.C. and S.S. supervised the project and acquired the funding. J.A.S. and T.S. prepared the manuscript, and all authors contributed to data interpretation, discussions, and writing.

## Acknowledgments

We thank Gaeun Kim of Prof. Yichun Wang’s lab, Department of Chemical and Biomolecular Engineering, University of Notre Dame, for providing the patient-derived healthy human dermal cell culture media for the initial experiments. We thank the Tissue Bank at the Harper Cancer Research Institute (HCRI), University of Notre Dame, for use of the NanoSight NS300 for nanoparticle tracking analysis. H.C.C. and S.S. acknowledge the partial support from the Common Fund through the Office of Strategic Coordination/Office of the NIH Director under 4UH3CA241684-03 and from the NIH, 1R21AI180713-01A1. Xin Lu acknowledges the support from NIH grants R01CA248033, R01CA280097, and R01CA297220.

## Supporting Information

**Supplementary Figure 1:**
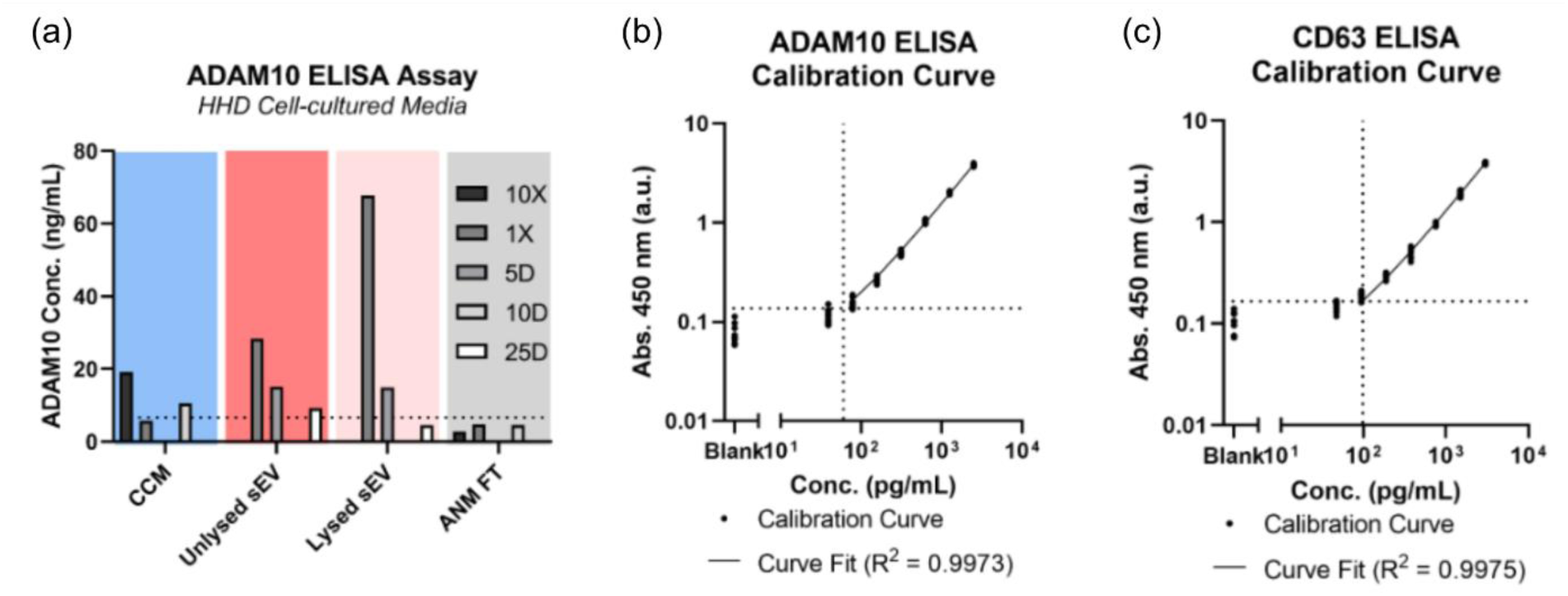
ELISA protein assays for CD63 and ADAM10. (a) ADAM10 ELISA assay results for healthy human dermal (HHD) cell-cultured media fractioned per workflow in Fig. 3a. CCM and FT fractions were tested in 10X concentrated using a 10kDa MWCO centrifugal filter and 1X & 10D dilutions in 1X DPBS. Unlysed and lysed sEV fractions were tested at 1X, 5D, and 25D dilutions in 1X DPBS. (b) Total ADAM10 ELISA calibration curve prepared per manufacturer’s instructions, featuring linear log-log fit (𝑅^2^=0.9973) between standard ADAM10 concentration and endpoint fluorescence assay after 20-minute incubation with a limit of detection of 0.14 FI corresponding to 60 pg/mL total ADAM10. (c) Total CD63 ELISA calibration curve prepared per manufacturer’s instructions, featuring a linear log-log fit (𝑅^2^=0.9975) between standard CD63 concentration and endpoint fluorescence assay after 20-min incubation with a limit of detection of 0.17 FI corresponding to 100 pg/mL total CD63.

**Supplementary Figure 2:**
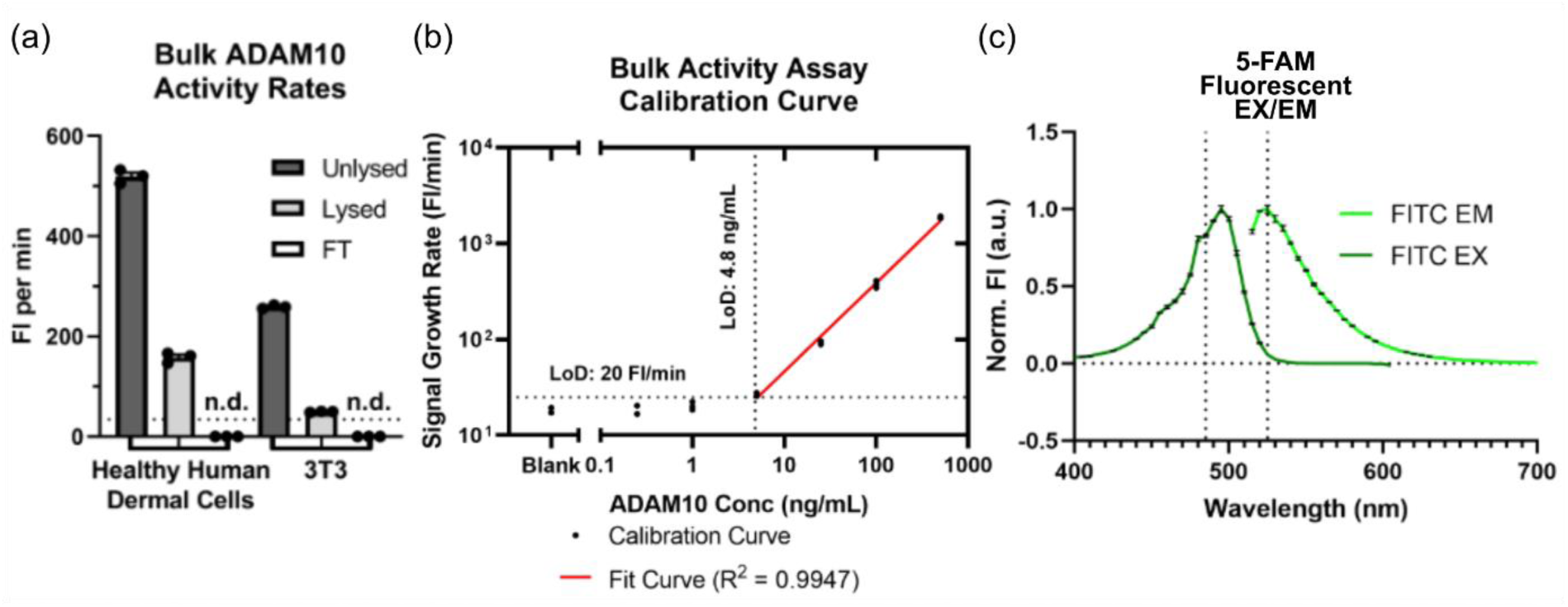
Bulk ADAM10 activity assays on additional cell lines. (a) Bulk ADAM10 activity rates for healthy human dermal (HHD) and healthy mouse fibroblast (3T3) cell-cultured media fractioned per workflow in Fig. 3a. (b) ADAM-10 bulk activity assay calibration curve prepared using serial dilutions of recombinant human ADAM-10 (rhADAM-10), featuring a linear log-log fit (𝑅^2^=0.9947) between rhADAM10 concentration and fluorescent signal growth rate with a limit of detection signal of 20 FI/min corresponding to 4.8 ng/mL rhADAM-10. (c) Excitation & emission spectra of 5-FAM fluorophore measured at fixed wavelengths. To measure excitation, a fixed emission wavelength was set to 525±20 nm. To measure emission, a fixed excitation wavelength was set to 485±10 nm.

**Supplementary Figure 3:**
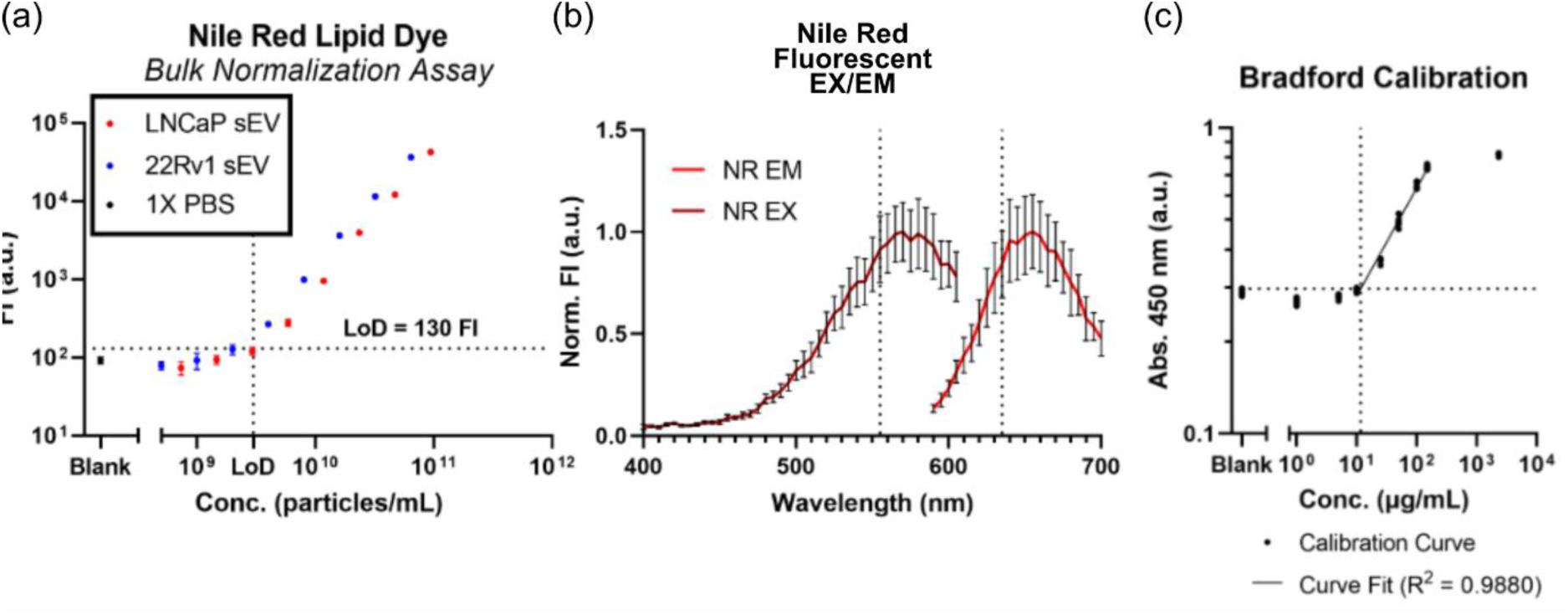
Additional information on Bradford total protein and Nile Red total lipid assays. (a) Nile Red fluorescent total lipid assay calibration curve prepared using serial dilutions of spin-filter isolated EVs from LNCaP and 22Rv1 cell cultured media, featuring a linear log-log fit (𝑅^2^=0.9960) between NTA-derived particle concentration and fluorescent intensities, with a limit of detection of 130 FI corresponding to 10^7^ particles/mL. (b) Excitation & emission spectra of Nile Red lipid dye incubated with diluted healthy human plasma measured at fixed wavelengths. To measure excitation, a fixed emission wavelength was set to 635±20 nm. To measure emission, a fixed excitation wavelength was set to 555±10 nm. (c) Bradford colorimetric total protein assay calibration curve prepared using serial dilutions of bovine serum albumin (BSA), featuring a linear log-log fit (𝑅^2^=0.9880) between protein concentration and absorbance values with a limit of detection of 0.30 FI corresponding to 12 μg/mL total protein.

